# SymPerturb converts symptom-network structure into testable intervention priorities

**DOI:** 10.64898/2026.07.27.26359002

**Authors:** Zheng Zhu, Junwen Yu, Tiantian Hu, Zhongfang Yang, Jiaqing Wang

**Affiliations:** School of Nursing, Fudan University, Shanghai, China; NYU Shanghai, Shanghai, China; School of Nursing, Soochow University, Suzhou, China; Yulin AI-Enhanced HealthCare Lab, Shanghai, China

## Abstract

Symptom networks encode conditional dependence but do not by themselves identify causal or clinically actionable intervention targets. We introduce SymPerturb, a virtual-perturbation framework that distinguishes four primitive perturbation operators—virtual knockout, virtual knockdown, edge-level communication blocking and node-centred communication blocking—from three analytic procedures—virtual dosage perturbation, combination perturbation and sequence optimisation. The reference Gaussian implementation is embedded in a general location–scale map with symptom-specific target anchors, making explicit that zero anchoring and linked mean–variance attenuation are modelling choices. Seven utility outcomes quantify downstream efficacy, dose efficiency, breadth, cross-module reach, communication blocking, combination value and responsiveness; robustness is reported separately as an uncertainty diagnostic. Their direction-aligned, within-candidate-set weighted mean defines the virtual perturbation priority score (VPPS), which is a relative ranking rather than a transportable clinical utility score. In a known 22-node, four-module generating network, analytical efficacy agreed with 100,000-draw Monte Carlo estimates within 0.0024 standard deviations. The reported finite-sample VPPS results were generated with the original eight-component exploratory score and therefore require regeneration under the revised seven-utility-dimension definition. These simulations provide internal computational verification under model compatibility, not causal or external validation. SymPerturb is intended to generate auditable target hypotheses for longitudinal and experimental testing.

## 1 Introduction

Network models have changed the way multivariate symptom data are represented. Instead of treating symptoms as interchangeable indicators of one latent disease, a symptom network represents them as interacting components whose conditional dependence may be summarised by a Gaussian graphical model (GGM) [1–4]. This representation has encouraged the use of strength, expected influence, bridge centrality and predictability to identify symptoms that appear structurally important [5–7]. Yet structural importance is not the same quantity as intervention value. A symptom may be highly connected because it is downstream of several common causes, may have strong but harmful and beneficial links that cancel, or may occupy a central position only within a particular choice of network boundary [8–10].

The gap becomes explicit when the scientific question changes from description to a model-based perturbation query. Descriptive centrality asks where a node sits in a fitted graph. A perturbation query asks how a prespecified fitted distribution or graph functional changes under an explicit operator. It does not ask what would happen under a real treatment unless the intervention semantics, temporal ordering and causal assumptions are independently justified. The query therefore requires three objects that centrality does not supply: a perturbation operator, a rule for updating the fitted system and a clinically interpretable outcome functional. Simulation-based intervention analysis and network-control approaches have begun to provide these objects [11–13], but their interpretation remains conditional on the generating model. In particular, conditioning on or statistically shifting a symptom in a cross-sectional undirected network is not, by itself, a causal do-intervention [14].

Here we formalise SymPerturb, a symptom-network virtual-perturbation framework, and retain the seven component names used in the original implementation. These components are not all objects of the same mathematical type. Virtual knockout, virtual knockdown, edge-level communication blocking and node-centred communication blocking are primitive perturbation operators. Virtual dosage perturbation is an intensity–response evaluation, combination perturbation is a joint-target construction and sequence optimisation is a decision procedure over ordered target sets. We also make three assumptions explicit. First, the zero endpoint is meaningful only after symptom orientation and clinical anchoring. Second, the reference state operator links mean attenuation to scale attenuation, which is a modelling choice rather than a necessary feature of symptom improvement. Third, exact knockout should be evaluated on a prespecified estimand rather than by re-estimating a full covariance matrix that retains a constant column.

SymPerturb is intended as a methods contribution. We derive the reference state update from a Gaussian regression decomposition, express topology blocking as matrix operators, define seven utility outcomes and one uncertainty diagnostic, and evaluate finite-sample recovery using data generated from a known network. The simulation is designed for internal computational verification: whether analytical and sampled responses agree, whether population rankings are recovered under model compatibility, how rankings change across analysis settings and when exact knockout breaks a downstream estimator. It does not establish external validity, clinical efficacy or causal transportability. Any ranking remains a model-derived hypothesis requiring longitudinal, experimental and patient-centred validation.

## 2 The SymPerturb architecture

SymPerturb contains a state layer, a topology layer and a multi-target layer (Fig. 1). The state layer contains two primitive operators: virtual knockout (vKO), which anchors a target to a prespecified state and may remove its residual variance, and virtual knockdown (vKD), which produces a partial location–scale change. Virtual dosage perturbation (vDP) evaluates an intensity–response curve generated by a state operator. The topology layer contains edge-level communication blocking and node-centred communication blocking, which attenuate one edge or all edges incident to a target, respectively. The multi-target layer contains combination perturbation and sequence optimisation; the latter searches ordered target sets under an explicit decision objective rather than inferring biological time from a cross-sectional network.

**Figure 1:**
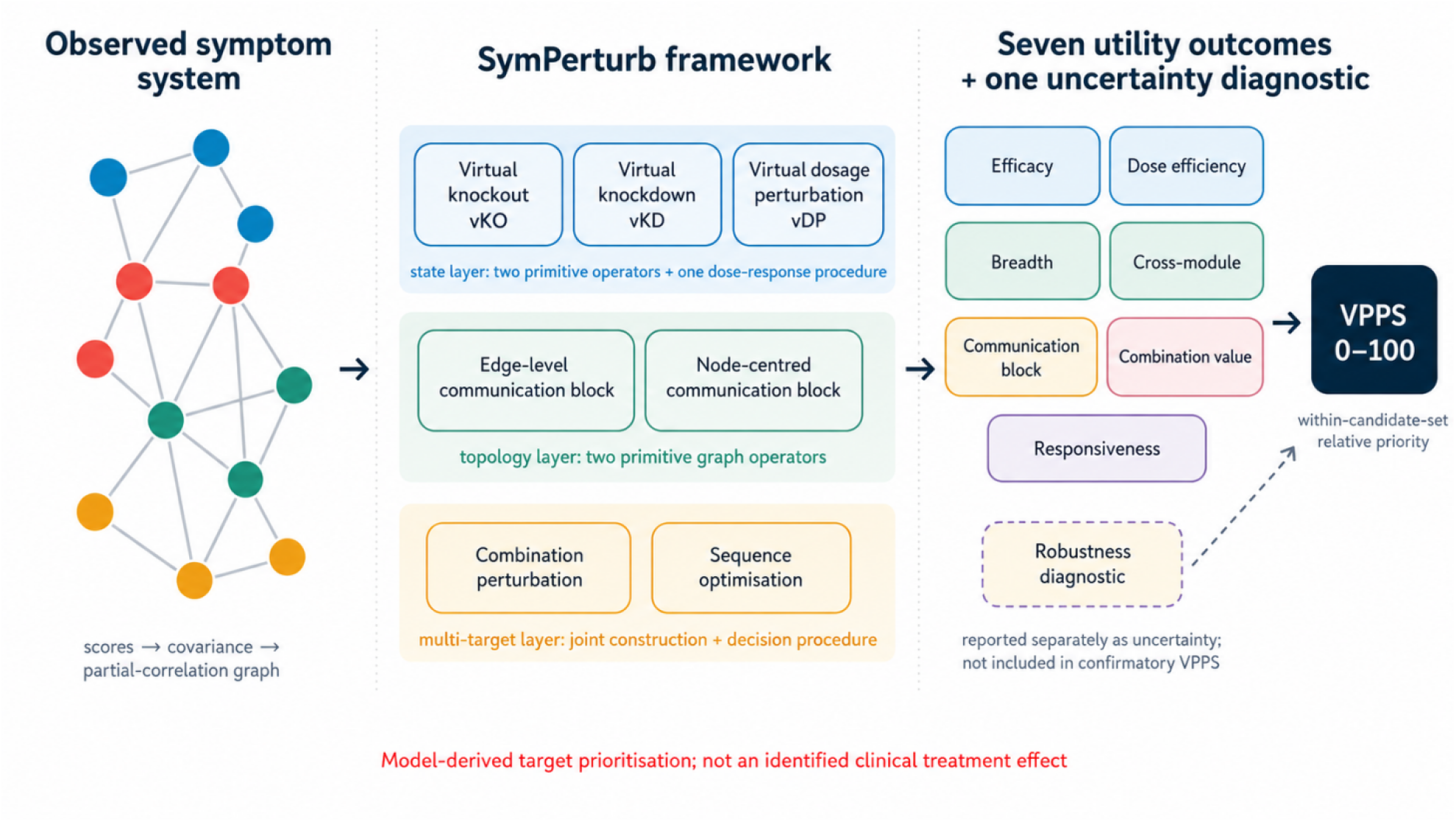
The SymPerturb workflow. Virtual perturbation specifies what is changed, how the fitted system is updated and how the response is evaluated. The original component names are retained, but their mathematical roles are distinguished: vKO and vKD are state operators; vDP is an intensity–response evaluation; edge-level and node-centred communication blocking are topology operators; combination perturbation is a joint-target construction; and sequence optimisation is a decision procedure. VPPS ranks model-derived target hypotheses within the analysed candidate set; robustness is reported separately, and neither quantity is an identified clinical treatment effect.

Each primitive operator returns either a distributional change or a topological change. The three analytic procedures evaluate, combine or optimise those operators. Seven utility outcomes then reduce the resulting changes to target-level quantities, while robustness is retained as a separate uncertainty diagnostic. VPPS is calculated only after the utility outcomes have been direction-aligned and normalised across the prespecified candidate set. This taxonomy prevents a category error: an edge weight, a model-implied symptom shift, a stable rank and an optimisation policy are different quantities. It also avoids mechanistic overstatement: “communication blocking” is retained as the framework label but denotes a graph attenuation operation, not evidence that symptoms literally exchange a biological signal.

## 3 A general location–scale state intervention with knockout as an endpoint

Let the symptom vector be partitioned into a target set S and the remaining symptoms K. Under a multivariate Gaussian model, the non-target vector has the regression representation

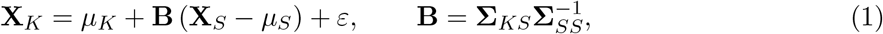

where

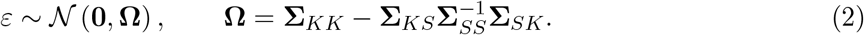

The matrix **Ω** is the Schur complement of **Σ**_*SS*_. A general state intervention may use symptom-specific target anchors and separate mean and scale maps, as formalised in Methods. The numerical results reported here use the reference special case in which all scales are oriented so that larger values indicate worse symptoms, the target anchor is zero, and the target mean displacement and standard deviation are both multiplied by **D**_*α*_ = diag(1 *− α*_*s*_). Under that linked special case, replace the target distribution by

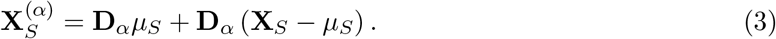

The intervention is exactly the baseline model when every *α*_*s*_ = 0. In the linked reference case, the target mean displacement from zero and the target standard deviation both shrink with dose, and the operator becomes an exact zero-anchored knockout when every *α*_*s*_ = 1. Substitution into the Gaussian regression representation gives

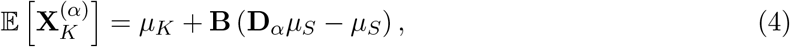

and

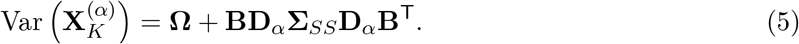

This linked location–scale formulation corrects a discontinuity in a simple hard-conditioning implementation: if a partial dose fixes a target to a constant, its variance collapses at any non-zero dose. However, linking mean attenuation and variance attenuation is a strong modelling assumption rather than a necessary property of treatment. SymPerturb therefore treats the linked map as a reference implementation and requires sensitivity analyses using location-only, scale-only and independently parameterised location–scale maps. It remains a statistical intervention defined within the fitted Gaussian model, not a causal intervention identified from cross-sectional data.

## 4 Seven components of SymPerturb

The seven components are summarised in Table 1 and derived in Methods. Four are primitive perturbation operators, whereas vDP, combination perturbation and sequence optimisation are an evaluation, a joint-target construction and a decision procedure, respectively. Virtual knockout is stated separately because its numerical consequences differ from those of partial knockdown. For a single target j, the unit-dose reference operator has

**Table 1:** Seven components of SymPerturb. Virtual knockout, virtual knockdown, edge-level communication blocking and node-centred communication blocking are primitive operators. Virtual dosage perturbation, combination perturbation and sequence optimisation are an intensity–response evaluation, a joint-target construction and a decision procedure, respectively.

| Component | Reference mathematical action | Mathematical role and primary question |
| --- | --- | --- |
| Virtual knockout, vKO | Exact reference case: $\alpha_j = 1$, target anchored at $c_j$; $c_j = 0$ and zero residual variance in the reported simulation | Primitive state operator: what is the model-implied response to exact target anchoring? |
| Virtual knockdown, vKD | $0 < \alpha_j < 1$; partially attenuate target location and scale under a prespecified map | Primitive state operator: what response follows a feasible partial target change? |
| Virtual dosage perturbation, vDP | Evaluate $G_j(\alpha)$ over $\alpha \in [0, 1]$ for a selected state operator | Intensity–response procedure: is the response linear, thresholded or saturating? |
| Edge-level communication block | $w'_{uv} = (1 - q)w_{uv}$ | Primitive topology operator: how does a selected graph functional change when one edge is attenuated? |
| Node-centred communication block | $w'_{jk} = (1 - q)w_{jk}$ for every $k \neq j$ | Primitive topology operator: how dependent is the selected graph functional on edges incident to one node? |
| Combination perturbation | Apply a joint operator to $\{j, k\}$ and compare with single-target operators on a common outcome set | Joint-target construction: does a pair add signed value beyond the better single target? |
| Sequence optimisation | Maximise discounted marginal gains over ordered target sets under explicit costs and constraints | Decision procedure: which feasible order maximises the prespecified objective? |

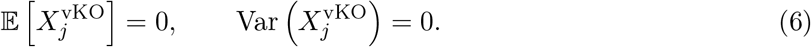

The non-target mean shift is

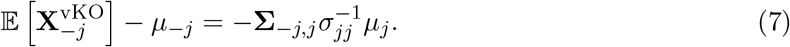

The direct implication is computational, not merely conceptual. A post-knockout data matrix containing a fixed target has a zero-variance column. Standardising that column is undefined and the full covariance matrix is singular. Network comparisons after exact vKO must therefore remove the target, treat its incident edges as absent, or use a pre-specified soft vKO with a small non-zero variance. These alternatives answer different questions and should not be interchanged without sensitivity analysis. In applied data, the knockout anchor must also be clinically justified; it need not equal zero.

## 5 Seven utility outcomes, a robustness diagnostic and VPPS

For target *j* at dose *α*, let 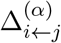 be the standardised improvement in non-target symptom i, and define the weighted system benefit

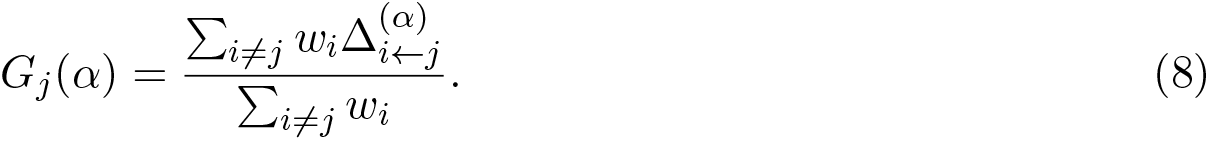

The default analysis used equal positive symptom weights. Higher values were oriented to indicate greater potential value for each utility outcome. Table 2 defines seven utility outcomes and one uncertainty diagnostic, with denominators, thresholds and common outcome sets made explicit. Robustness is not a clinical utility dimension and is therefore excluded from the confirmatory VPPS.

**Table 2:** Seven utility outcomes and one robustness diagnostic in SymPerturb. *q* is the number of modules; *c*(*j*) is the module of target *j*; *Q*_*T*_ is a finite-step topology functional; and *T*_*j*_ is the pre-specified partner set. Robustness is reported separately and is not included in the confirmatory VPPS. The combination score is a model-based incremental value measure, not an additive causal interaction.

| Outcome | Definition | Interpretation |
| --- | --- | --- |
| Downstream efficacy score | Full-dose system benefit, $G_j(1)$ | Mean standardised non-target improvement after full vKO; direct target benefit is reported separately |
| Dose-efficiency score | Mean of $G_j(\alpha_k)/\alpha_k$ over the pre-specified doses | Benefit per unit intervention over partial doses |
| Breadth score | Share of non-target improvements satisfying $\Delta_{i \leftarrow j}^{(1)} \geq \tau$ | Proportion of non-target symptoms exceeding an improvement threshold |
| Cross-module score | Share of other modules with mean improvement at least $\tau_m$ | Proportion of other symptom modules reached |
| Robustness diagnostic | One minus the normalised standard deviation of scenario ranks | Sensitivity of target rank across scenarios; uncertainty diagnostic, not included in confirmatory VPPS |
| Communication-block score | Relative loss in $Q_T$ after node-centred blocking | Relative loss of a prespecified finite-step topology functional after node-centred blocking |
| Combination-value score | Mean positive incremental pair value over partner set $\mathcal{T}_j$ in the current exploratory implementation | Incremental value over the better single target; confirmatory analyses should retain signed values |
| Responsiveness score | $\{G_j(\epsilon) - G_j(0)\}/\epsilon$ | Local low-dose sensitivity of the system |

Each raw utility outcome *R*_*jm*_ is normalised across the prespecified candidate targets:

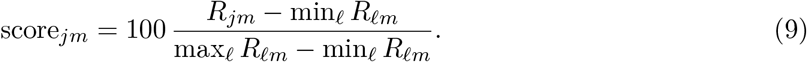

If a utility dimension is constant, it contributes a neutral score of 50 rather than an undefined value. The confirmatory VPPS is the weighted mean of the seven utility dimensions:

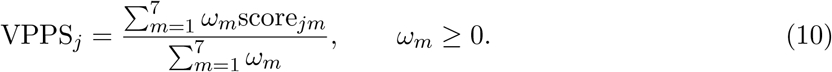

We used equal weights, *ω*_*m*_ = 1, for the reference methodological analysis. These weights are not clinical utilities. A clinical application should pre-specify weights using patient priorities, intervention feasibility, safety and decision-analytic considerations, then report the unaggregated seven-dimensional utility profile and the separate robustness diagnostic beside VPPS. Because min–max normalisation is performed within the candidate set, VPPS is a relative within-analysis ranking and cannot be directly compared across networks, cohorts or candidate sets. The numerical VPPS results currently reported in this draft were generated with the original eight-component exploratory composite and must be regenerated under the revised seven-dimension confirmatory rule.

## 6 Internal computational verification and finite-sample recovery

We generated a 22-node GGM with four symptom modules, positive and negative edges, heterogeneous means and heterogeneous variances. The precision matrix was positive definite by construction and the observed 0–4 scores were obtained by boundary clipping. Population outcomes were calculated from the known generating parameters. At each of n=250, 500 and 1,000, 200 independent datasets were generated, fitted and ranked. The independently generated dataset, not the node or Monte Carlo draw, was the statistical unit. This design provides internal verification under model compatibility; it does not test model misspecification or external validity.

Analytical vKO efficacy closely matched direct simulation. Across 22 targets and 100,000 conditional draws per target, the maximum absolute discrepancy was 0.0024 standard deviations (Fig. 2a). This verifies the numerical implementation of the linked reference map and boundary-expectation calculations. It does not verify the clinical truth of the GGM, the target anchor or the linked mean–scale assumption.

**Figure 2:**
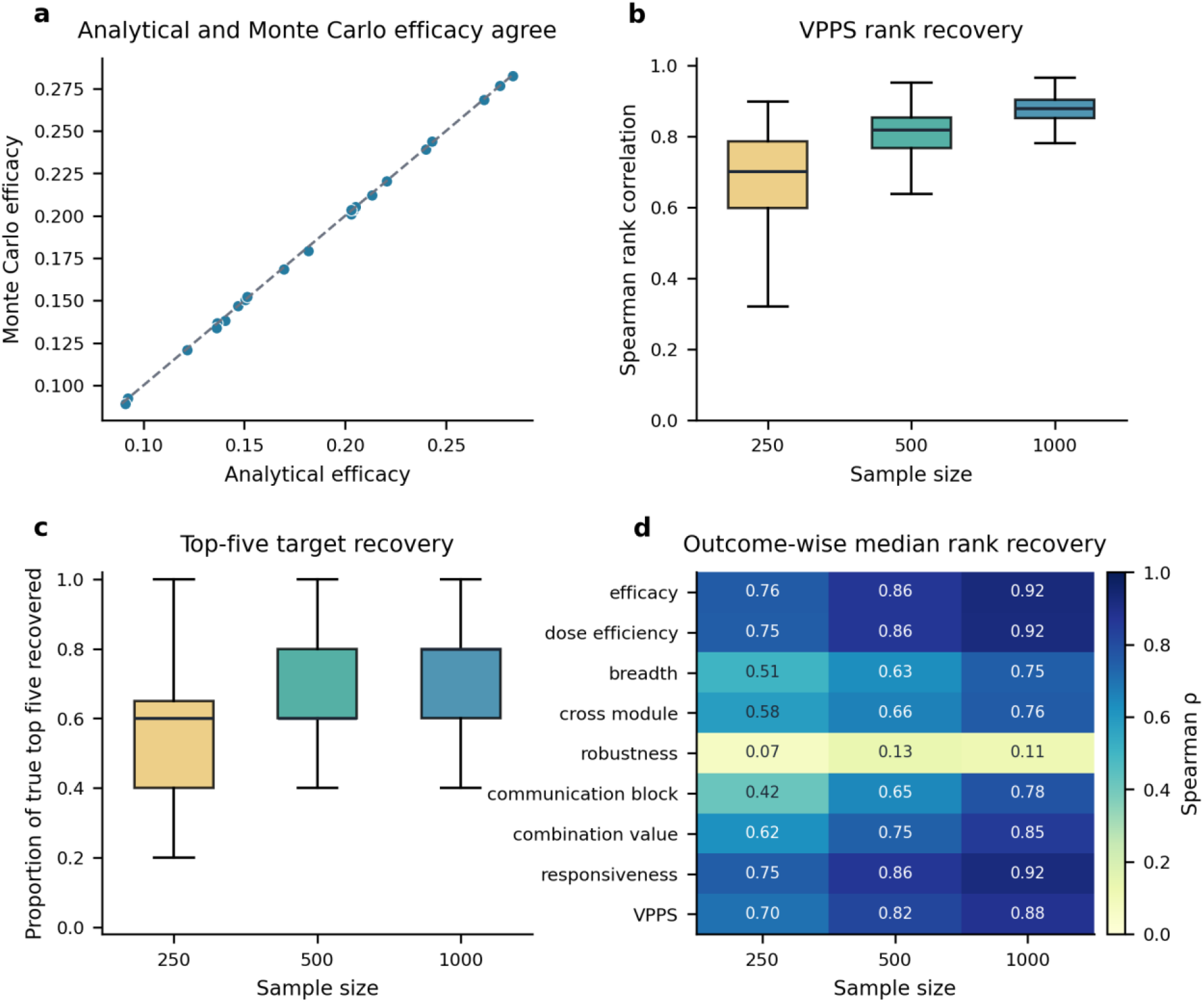
Recovery of perturbation outcomes in model-compatible simulated networks. (a) Analytical efficacy versus Monte Carlo efficacy for 22 targets, with the identity line. (b) Distribution of the original exploratory eight-component VPPS rank correlation across 200 independently generated datasets at each sample size. (c) Proportion of the population top five recovered. (d) Median outcome-specific rank correlation. These panels do not contain results for the revised seven-utility-dimension confirmatory VPPS, which requires reanalysis.

The original exploratory eight-component VPPS ranking improved consistently with sample size (Fig. 2b and Table 3). Median Spearman correlation with the population exploratory VPPS rank was 0.70 at n=250, 0.82 at n=500 and 0.88 at n=1,000. The median proportion of true top-five targets recovered was 0.60, 0.60 and 0.80, respectively (Fig. 2c). Efficacy, dose efficiency and responsiveness had the strongest rank recovery. Communication blocking and threshold-based breadth and cross-module scores required larger samples. The robustness diagnostic was the clear exception: median rank recovery was 0.07, 0.13 and 0.11 across the three sample sizes (Fig. 2d). These numerical results are retained as an audit trail but must be regenerated after robustness is removed from the confirmatory VPPS.

**Table 3:** Finite-sample recovery of the original exploratory eight-component VPPS. Intervals are the 2.5th and 97.5th percentiles across 200 independently generated datasets, not confidence intervals for a patient population. The table must be regenerated for the revised confirmatory VPPS.

| Sample size | Median exploratory VPPS8 $\rho$ (95% Monte Carlo interval) | Median top-five recovery (95% interval) | Median exploratory VPPS8 RMSE on 0–100 scale (95% interval) |
| --- | --- | --- | --- |
| 250 | 0.70 (0.39–0.88) | 0.60 (0.20–0.80) | 16.35 (11.17–23.20) |
| 500 | 0.82 (0.65–0.92) | 0.60 (0.40–1.00) | 13.18 (9.47–17.40) |
| 1,000 | 0.88 (0.80–0.94) | 0.80 (0.40–1.00) | 10.58 (7.86–13.97) |

## 7 Knockout validity, sensitivity and distinction from benchmark rankings

Exact vKO exposed a deterministic failure mode. The target column had zero variance in every one of 100 diagnostic replicates; consequently, full-network standardisation and covariance re-estimation were invalid in all replicates. Deleting the target and estimating the induced subgraph was valid in all 100 replicates, as was a soft vKO retaining 5% of the target standard deviation (Fig. 3b). An exact knockout result should therefore report the target anchor and whether it represents state anchoring, graph deletion or a soft intervention.

**Figure 3:**
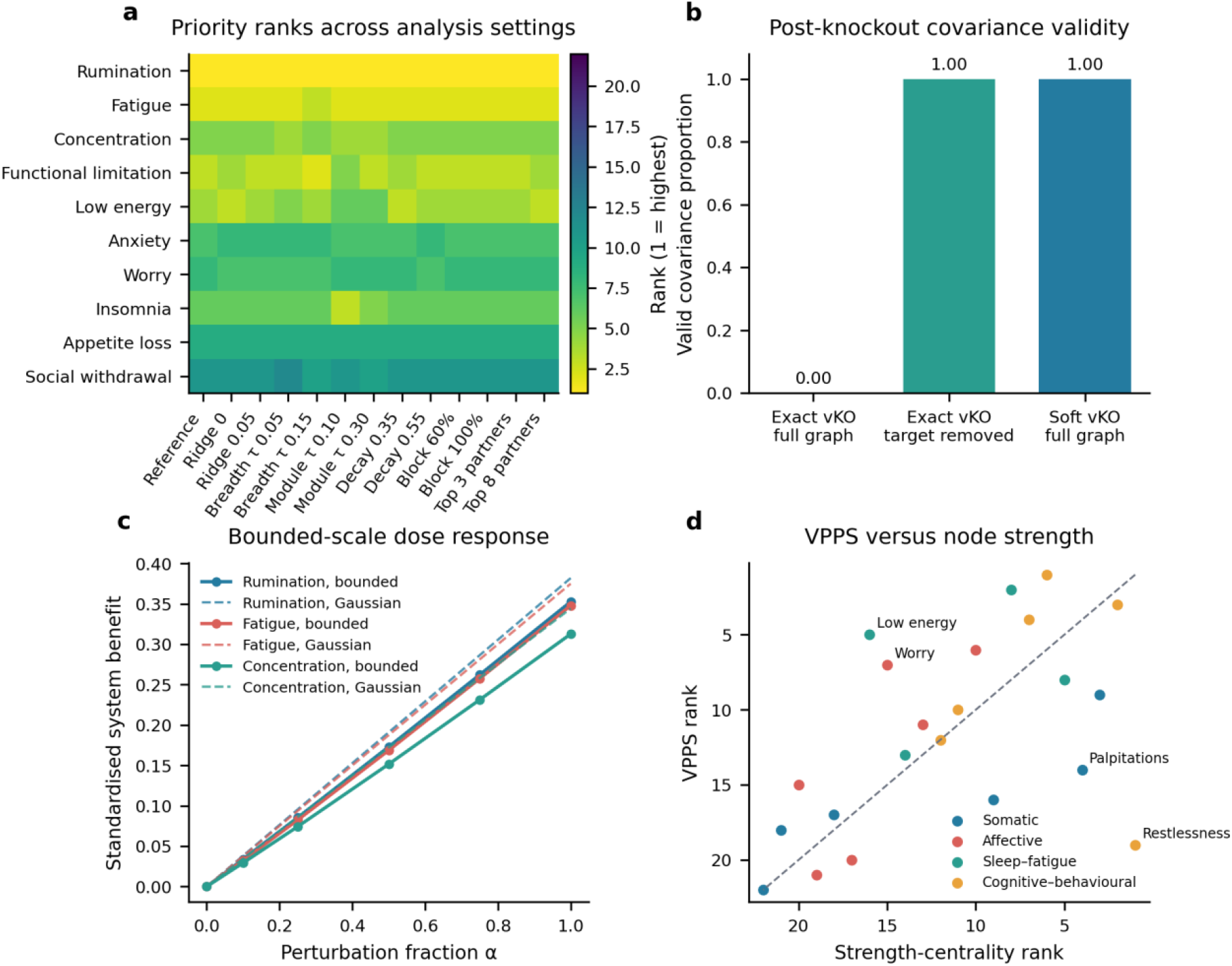
Robustness, boundary effects and the centrality contrast. (a) Population target ranks under 13 analysis scenarios. (b) Valid covariance proportion across 100 exact, reduced-network and soft-knockout replicates. (c) Bounded and unbounded dose curves in a high-burden saturation stress test. (d) Population exploratory VPPS rank versus absolute strength-centrality rank; colours denote symptom modules. The strength comparison is illustrative rather than a complete benchmark.

The population exploratory ranking was insensitive to the 13 pre-specified analysis scenarios. Their Spearman correlations with the reference provisional ranking had a median of 0.998 and ranged from 0.983 to 1.000 (Fig. 3a). This high population stability did not make the estimated robustness diagnostic reliable, because small sample changes could still rearrange near-tied scenario ranks. Population sensitivity and sampling uncertainty are distinct and should be reported separately through scenario profiles, bootstrap rank distributions and top-k selection probabilities.

The 0–4 boundary produced visible curvature in a high-burden saturation stress test, whereas the corresponding unbounded Gaussian response was linear (Fig. 3c). Thus efficacy, dose efficiency and responsiveness can be almost redundant under a linear Gaussian model but diverge under range restriction or other non-linear measurement functions. Their empirical correlation should be inspected before they are combined, and sensitivity analyses should compare location-only, scale-only and linked state maps.

Finally, the population exploratory VPPS rank was only moderately associated with absolute strength-centrality rank, *ρ*=0.52 (Fig. 3d). In this synthetic example, Rumination, Fatigue, Concentration, Functional limitation and Low energy had the highest population values. These names are labels in generated data, not clinical recommendations. The contrast shows that SymPerturb differs from node strength, but comparison with strength alone is insufficient to establish incremental value. A confirmatory benchmark should include expected influence, bridge metrics, predictability, control-based metrics and a burden-only rule under pre-specified ranking, top-k and decision-regret losses.

## 8 Discussion

SymPerturb converts a descriptive symptom network into a set of explicit, auditable target hypotheses. Its main contribution is not another centrality statistic. It separates primitive perturbation operators from the procedures that evaluate, combine or optimise them, and separates clinical-utility dimensions from estimation uncertainty. Virtual knockout and knockdown act on symptom distributions; communication blocking acts on graph topology; virtual dosage perturbation evaluates an intensity–response curve; combination perturbation constructs joint targets; and sequence optimisation searches an ordered decision objective. Seven utility outcomes answer questions about magnitude, efficiency, reach, topology dependence, partner value and local sensitivity, while robustness characterises uncertainty rather than value.

The mathematical formulation clarifies several quantities that can otherwise be overinterpreted. First, exact vKO is not simply “100% knockdown” for all downstream computations. It is a degenerate endpoint at a prespecified anchor and invalidates a full-network covariance estimate when the target variance is zero. Second, the reference map assumes that mean displacement and standard deviation contract together; location-only, scale-only and independently parameterised maps may represent different intervention semantics. Third, dose efficiency and responsiveness do not automatically add independent information. In an unbounded linear Gaussian model, *G*_*j*_(*α*) = *αG*_*j*_(1), so efficacy, per-dose benefit and low-dose slope are algebraically equivalent. Fourth, the communication-block score is a property of the chosen topology functional, including edge thresholding, sign handling, scaling, decay and path length. It is not direct evidence that symptoms exchange a biological signal.

The validation provides both support and restraint. Analytical and Monte Carlo state responses agreed to within 0.0024 standard deviations, and recovery of the original exploratory VPPS improved with sample size. Nevertheless, at n=250, the 95% Monte Carlo interval for rank recovery extended from 0.39 to 0.88, and top-five recovery could be as low as 0.20. More importantly, the rank-standard-deviation robustness diagnostic was poorly recovered even at n=1,000. Robustness should therefore be reported through scenario-specific ranks, bootstrap selection probabilities and uncertainty intervals rather than included in VPPS. All composite results must be regenerated using the revised seven-utility-dimension definition.

Several limitations define the scope of the claims. The current verification uses one family of synthetic GGMs and an approximately continuous 0–4 symptom scale. It does not cover strongly ordinal or zero-inflated data, latent confounding, measurement error, missingness, longitudinal or person-specific systems, estimator misspecification, or high-dimensional p/n regimes. The thresholds for breadth and module reach are analysis choices, not minimal clinically important differences. Equal VPPS weights are a methodological convention, not a clinical preference model. The combination score measures improvement beyond the better single target and, in its current positive-part form, suppresses harmful joint effects; a confirmatory implementation should retain signed incremental values. Sequence optimisation uses a discounted set objective and a restricted candidate search; it does not solve a general dynamic treatment regime. Most importantly, a cross-sectional undirected GGM does not identify intervention direction or remove unmeasured confounding [8–10, 14].

A credible translational pathway should therefore proceed in stages. The first stage is expanded computational validation, including factorial model-misspecification simulations, complete-pipeline bootstrap uncertainty, parameter sensitivity, systematic benchmark comparisons and independent software replication. The second is longitudinal validation, asking whether natural or treatment-induced changes in a candidate target precede the predicted downstream changes. The third is experimental validation with measured target engagement, ideally followed by prospective decision studies that incorporate safety, cost, equity and patient preferences. Used within these boundaries, SymPerturb can narrow a target set and make assumptions testable. It cannot replace a trial.

## 9 Methods

### 9.1 Baseline Gaussian graphical model

Let **X** ∈ ℝ^*p*^ denote continuous or approximately continuous symptom scores:

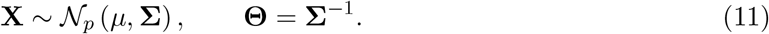

The partial correlation between symptoms i and j, conditional on the other symptoms, is

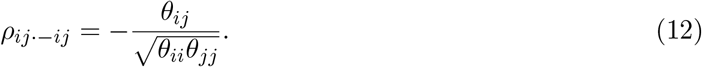

For finite-sample estimation, the empirical covariance S was diagonally regularised:

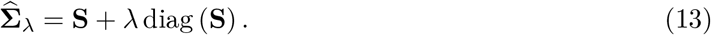

The simulation used *λ* = 0.02. Partial correlations smaller than *τ*_*W*_ = 0.03 in absolute magnitude were set to zero for topology-based outcomes. This is ridge-like covariance regularisation followed by hard edge thresholding, not graphical LASSO. Thresholded partial correlations were used as an adjacency matrix W; they were not assumed automatically to define a new positive-definite precision matrix. Because topology outcomes depend strongly on regularisation and thresholding, applied analyses should propagate network-estimation uncertainty through the complete estimation– perturbation–scoring pipeline rather than relying only on a small set of deterministic sensitivity scenarios.

### 9.2 SymPerturb operator taxonomy, target anchors and estimands

SymPerturb distinguishes primitive perturbation operators from analytic procedures. Virtual knockout, virtual knockdown, edge-level communication blocking and node-centred communication blocking are primitive operators. Virtual dosage perturbation evaluates an intensity–response curve, combination perturbation constructs a joint target and sequence optimisation searches an ordered decision objective. All symptom variables should be oriented so that larger values represent worse states. Let **c**_*S*_ denote clinically interpretable, symptom-specific target anchors, which need not equal zero. A general location–scale map is

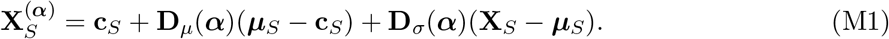

The corresponding target mean and covariance are **c**_*S*_ + **D**_*µ*_(***α***)(***µ***_*S*_ *−* **c**_*S*_) and **D**_*σ*_(***α***)**Σ**_*SS*_**D**_*σ*_(***α***), respectively. Baseline recovery requires **D**_*µ*_(0) = **D**_*σ*_(0) = **I**, whereas an exact knockout at the anchor requires **D**_*µ*_(1) = **D**_*σ*_(1) = **0**. The present numerical implementation uses the linked special case **c**_*S*_ = **0** and **D**_*µ*_(*α*) = **D**_*σ*_(*α*) = **D**_*α*_. Sensitivity analyses should compare location-only attenuation (**D**_*σ*_ = **I**), scale-only attenuation (**D**_*µ*_ = **I**) and independently parameterised location–scale maps.

Three estimands should be distinguished. Direct target benefit evaluates change in the perturbed targets S. Beneficial downstream spillover evaluates improvement in K, whereas adverse spillover records worsening in K. The current *G*_*j*_(*α*) is a downstream-spillover estimand and therefore does not represent total clinical benefit.

### 9.3 Derivation of the location-scale state intervention

Partition X, *µ* and Σ into targets S and non-targets K. The Gaussian conditional mean is

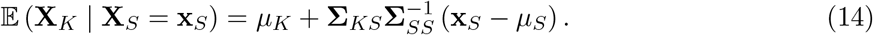

Define

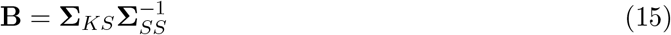

and the residual

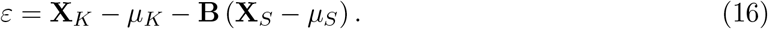

Its cross-covariance with the targets is zero:

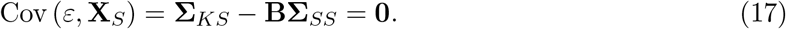

Joint normality therefore implies independence, and the residual covariance is the Schur complement Ω defined in the Main text. The intervention replaces the target component by

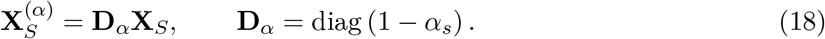

This compact form is equivalent to the centred, zero-anchor linked special case in the Main text because **D**_*α*_**X**_*S*_ = **D**_*α*_***µ***_*S*_ + **D**_*α*_(**X**_*S*_ *−* ***µ***_*S*_). Substitution gives the non-target mean and covariance shown above. The full post-intervention cross-covariance is

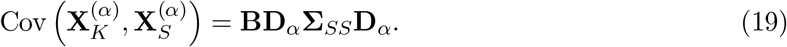

At zero dose, **D**_*α*_ = **I** and all baseline moments are recovered. At unit dose, **D**_*α*_ = **0**, the target becomes a constant at the zero anchor and the non-target covariance reduces to the Schur complement. These statements describe the linked reference map; the general anchor and separate location–scale maps are defined above.

### 9.4 Bounded symptom scales

The primary simulation treated symptom scores as approximately continuous but restricted reported scores to [0, 4] using

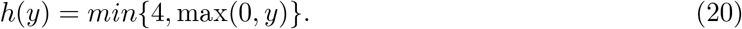

For *Y ∼N* (*m, − s*^2^), let *a* = (0 *− m*)*/s* and *b* = (4 *− m*)*/s*. Integration over the two tails and the interior gives

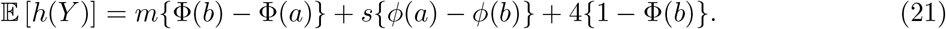

When s=0, the expectation is h(m). This expression was used to calculate population and fitted post-intervention means without Monte Carlo sampling. It is a winsorised-normal expectation, not a truncated-normal likelihood. The high-burden stress test increased means by 0.85, with a maximum of 3.75, to expose boundary-induced dose curvature.

### 9.5 Virtual knockout

For target j, vKO is the unit-dose state operator:

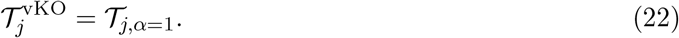

In the numerical reference implementation, the target distribution is degenerate at the zero anchor. State outcomes can be calculated analytically because only the target variance is zero. If a post-knockout GGM is required, the primary recommendation is to estimate the induced network on *V \*{*j*} . A topology-only knockout may instead set all *w*_*jk*_ = 0. A soft vKO can retain a pre-specified variance 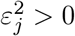, but *ε*_*j*_ and the target anchor *c*_*j*_ must be reported and varied in sensitivity analysis.

### 9.6 Virtual knockdown and dosage perturbation

Partial knockdown uses 0 *< α <* 1. For one target, the unbounded Gaussian mean response is

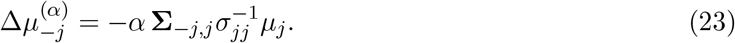

Consequently,

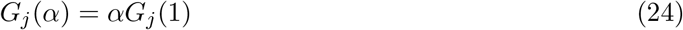

when the scale is unbounded, the covariance is fixed and the outcome is linear in the non-target mean changes under the linked reference map. Dose efficiency and responsiveness are then redundant with efficacy. The implemented dose grid was *α ∈* {0,0.10,0.25,0.50,0.75,1}. Departure from linearity was evaluated by comparing the bounded response with the corresponding unbounded Gaussian response. Confirmatory analyses should additionally compare location-only attenuation, scale-only attenuation and the linked map.

### 9.7 Edge-level and node-centred communication blocking

For an undirected edge (u,v) with block fraction *q ∈* [0, 1],

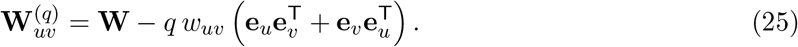

Node-centred blocking attenuates all edges incident to j:

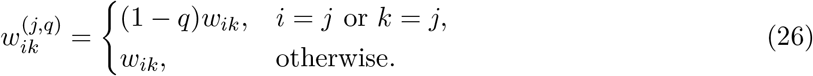

The reference block fraction was q=0.80. Because these operators act directly on W, they evaluate graph topology unless the modified graph is converted back to a valid precision matrix. Such coupling would require preserving conditional-variance scales, enforcing positive definiteness and re-inverting the precision matrix. The term “communication” is therefore shorthand for topology attenuation and should not be read as a biological transmission mechanism.

Finite-step propagation potential was defined as

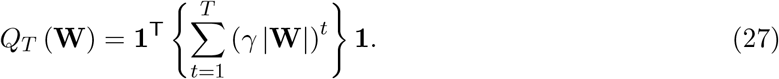

The simulation used T=6 and *γ*=0.45. Absolute weights measure unsigned route capacity irrespective of sign. Although finite T avoids a formal infinite-series convergence requirement, the magnitude of *Q*_*T*_ can still be dominated by node degree, cycles and matrix scaling. A confirmatory implementation should report the spectral radius of *γ* |**W**|, compare raw, row-normalised and spectral-normalised adjacency matrices, and report signed and unsigned propagation functionals when inhibitory relations are scientifically meaningful.

### 9.8 Combination perturbation

For targets j and k, the joint operator applies unit dose to both targets using the multivariate location-scale equations. To avoid attributing direct removal of either target to one method but not the other, all pair comparisons use the common outcome set *V\*{ *j, k*} . Let *G*_*jk*_(1, 1) be the mean standardised improvement on that set, and let 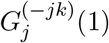 and 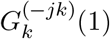 be the corresponding single-target benefits. Incremental pair value is

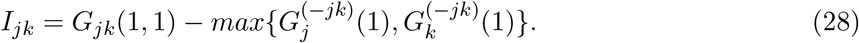

This contrast quantifies improvement beyond the better single target. It is deliberately not called additive synergy, which would instead compare the joint effect with the sum of single effects under an explicit causal interaction model. The current exploratory implementation averages only positive incremental values; confirmatory analyses should retain signed pair values so that antagonistic or harmful combinations remain visible. For each target, the reference partner set contained the five highest-efficacy alternative targets.

### 9.8 Sequence optimisation

Let ***π*** = (*π*_1_, …, *π*_*L*_) be an ordered sequence and *S*_*t*_ = {*π*_1_, …, *π*_*t*_}. A discounted sequence objective was

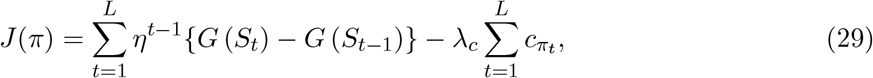

where 0 *< η* ≤ 1 favours earlier benefit and 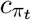 is intervention cost. Without discounting, order-dependent costs or constraints, or adaptive model updating, the objective is order invariant for a fixed final target set and reduces to subset selection. The sequence is therefore induced by the decision objective, feasibility constraints and costs—not by biological or temporal direction inferred from a cross-sectional GGM.

### 9.9 Seven utility outcomes and one robustness diagnostic

For equal weights, standardised improvement was

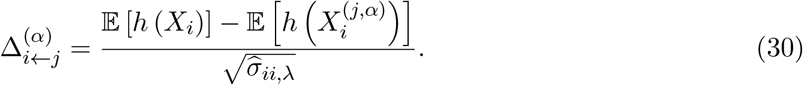

The target itself was excluded from every downstream system-benefit, breadth and single-versus-pair comparison. The current *G*_*j*_(*α*) is therefore a downstream-spillover estimand rather than total clinical benefit. Applied analyses should report direct target benefit, beneficial downstream spillover and adverse downstream spillover separately. Reference thresholds were *τ* =0.10 standard deviations for breadth and *τ*_*m*_ = 0.20 for a module-average improvement. Dose efficiency used *α*_*k*_ *∈* {0.25, 0.50, 0.75}, responsiveness used *ε* = 0.10, and communication blocking used the finite-step topology definition above.

The seven utility outcomes and the separate robustness diagnostic were calculated as

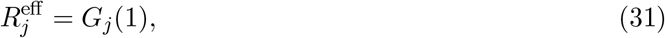

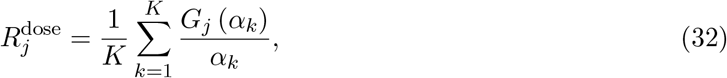

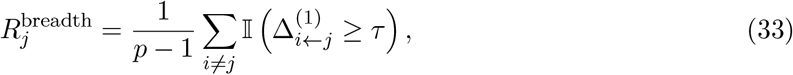

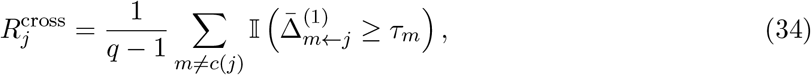

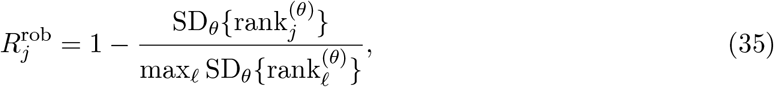

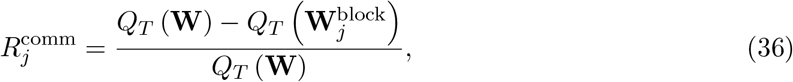

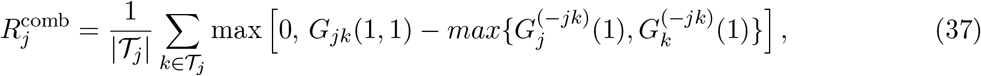

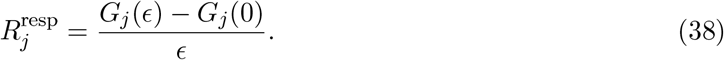

Robustness was calculated from a provisional composite of the other seven normalised outcomes. The 13 scenarios were: reference settings; ridge values 0 and 0.05; breadth thresholds 0.05 and 0.15; module thresholds 0.10 and 0.30; propagation decay 0.35 and 0.55; block fractions 0.60 and 1.00; and partner-set sizes 3 and 8. The standard deviation of each target rank across these scenarios was converted to the robustness diagnostic in Table 2. Because this quantity was weakly recoverable and reflects uncertainty rather than utility, it is excluded from the revised confirmatory VPPS. The numerical results in the current draft were generated before this revision and require reanalysis.

### 9.11 Simulation design

The generating network had 22 named symptom nodes and four modules: Somatic, Affective, Sleep–fatigue and Cognitive–behavioural. Within-module edges were sampled more densely and strongly than cross-module edges. Additional bridge edges connected Fatigue with Functional limitation and Concentration, Worry with Insomnia, Pain with Insomnia, Sadness with Social withdrawal, Anxiety with Palpitations, Low energy with Anhedonia and Rumination with Insomnia. A minority of edges were negative. The symmetric partial-correlation matrix was scaled to spectral radius at most 0.68, and

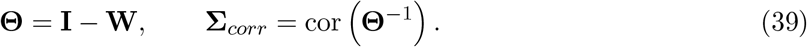

Node standard deviations ranged from 0.60 to 0.84 and means ranged from 1.30 to 2.80. The covariance was diag(**s**)**Σ**_corr_ diag(**s**). Each dataset was sampled from the resulting multivariate normal distribution and clipped to [0, 4]. The full parameter values and a generated n=1,000 dataset accompany the code.

Population outcomes were computed from the known generating parameters. At each sample size, 200 independent datasets were generated from distinct child seeds of the master seed 20260727. Sample means and covariances were then passed through the same fitting and scoring functions. The primary recovery statistic was Spearman correlation between estimated and population target ranks. We also calculated the proportion of the population top five recovered and root mean squared error of the original exploratory VPPS on its 0–100 scale. This design evaluates finite-sample recovery under correct model specification; it does not assess robustness to misspecification.

The analytical-versus-Monte Carlo check used 100,000 post-vKO draws for each of 22 targets. The vKO validity check used 100 independently generated post-intervention datasets of size 500 and compared exact full-network vKO, exact vKO with the target removed, and soft vKO retaining 5% of the target standard deviation. Parameter-sensitivity correlations compared each scenario with the reference population rank.

### 9.12 Required confirmatory validation extensions

The numerical results in this draft provide internal verification under a single model-compatible GGM family. A general methods claim requires a factorial stress-test suite that varies data type (continuous, ordinal and zero-inflated), latent confounding, measurement error, missingness, network density, edge-sign balance, module strength, p/n ratio, estimator and regularisation or thresholding strategy. Performance should be summarised across factors rather than within one favourable generating mechanism.

SymPerturb should be benchmarked against absolute strength, expected influence, bridge strength or bridge expected influence, predictability, control-based metrics and a burden-only rule. The comparison should pre-specify losses such as population-rank recovery, top-k recovery, expected decision regret, stability and calibration of predicted downstream change.

A complete-pipeline bootstrap should repeat network estimation, threshold selection, perturbation, outcome calculation, candidate-set normalisation and ranking. An empirical benchmark should then test whether longitudinal or experimentally induced target changes precede the predicted downstream changes and whether the intervention demonstrably engages the intended target.

### 9.13 Statistical analysis

No null-hypothesis significance tests were used. The independent unit for finite-sample verification was an independently generated dataset. Medians and 2.5th to 97.5th percentile Monte Carlo intervals summarised the 200 replicate distributions. These intervals quantify simulation-to-simulation variability under the specified generating model; they are not confidence intervals for patients, diseases or treatment effects. Monte Carlo draws used to verify an analytical expectation were not treated as additional independent datasets. For applied analyses, nonparametric or parametric bootstrap resampling should repeat network estimation, thresholding, perturbation, outcome calculation, normalisation and ranking in every replicate, yielding uncertainty intervals, rank distributions and top-k selection probabilities.

### 9.14 Software and reproducibility

The supplied R implementation defined the original symptom labels, module structure and conceptual perturbation workflow. The revised validation was executed in Python 3.12 using NumPy, pandas, SciPy and Matplotlib because an R runtime was not available in the execution environment. The accompanying script contains data generation, the original eight outcomes, VPPS calculation, validation and figure export. Before submission, the software must be updated to implement symptom-specific anchors, separate location and scale maps, signed combination values, a seven-utility-dimension VPPS, separate robustness diagnostics and complete-pipeline bootstrap uncertainty. An independently reviewed implementation in the intended analysis environment should reproduce all source-data tables.

Generative artificial intelligence was used to assist manuscript drafting, mathematical cross-checking and code review. All equations, generated data, numerical outputs and interpretations require author verification, and accountability remains with the named authors.

## Data Availability

All data produced in the present study are available upon reasonable request to the authors

## Data availability

All data used in this draft are synthetic. The generated n=1,000 example dataset, population parameters, source data underlying the validation results and target-level scores accompany the reproducibility bundle. Before submission, these files should be deposited in a public research repository with a persistent identifier. No patient data were used.

## Code availability

The core computational code underlying the original virtual-perturbation implementation has been published previously in a related methodological article. The present study extends that implementation by introducing the SymPerturb framework, including general target anchors, separate location–scale mappings, seven utility outcomes, robustness as a separate uncertainty diagnostic and full-pipeline uncertainty assessment. The revised code used for the analyses reported here, together with machine-readable outputs and source data, will be made available in a public, version-controlled repository upon publication. The previously published R code will be retained as methodological provenance, whereas the updated SymPerturb definitions described in this Article will govern the validated implementation.

## Acknowledgements

This work was supported by the National Natural Science Foundation of China (Grant No. 72574043), the Shanghai Pujiang Program (Grant No. 24PJC014), and the China University Industry–Research Innovation Fund—Digital Intelligence Innovation and Talent Program (Grant No. 2024LC007), all awarded to Z.Z. The funders had no role in the study design, data collection, data analysis, interpretation of the results, decision to publish, or preparation of the manuscript.

## Author contributions

Z.Z. conceived and developed the SymPerturb framework, designed the methodological study, supplied the original R implementation and evaluation specifications, and drafted the manuscript.

J.Y., T.H. and Z.Y. provided methodological and conceptual feedback and critically reviewed and revised the manuscript. All authors reviewed and approved the final version of the manuscript.

## References

[1] Denny Borsboom and Angelique O. J. Cramer. Network analysis: an integrative approach to the structure of psychopathology. Annual Review of Clinical Psychology, 9:91–121, 2013.

[2] Denny Borsboom et al. Network analysis of multivariate data in psychological science. Nature Reviews Methods Primers, 1:58, 2021.

[3] Sacha Epskamp, Denny Borsboom, and Eiko I. Fried. Estimating psychological networks and their accuracy: a tutorial paper. Behavior Research Methods, 50:195–212, 2018.

[4] Donald R. Williams and Philippe Rast. Back to the basics: rethinking partial correlation network methodology. British Journal of Mathematical and Statistical Psychology, 73:187–212, 2020.

[5] Donald J. Robinaugh, Alexander J. Millner, and Richard J. McNally. Identifying highly influential nodes in the complicated grief network. Journal of Abnormal Psychology, 125:747–757, 2016.

[6] Payton J. Jones, Ruijie Ma, and Richard J. McNally. Bridge centrality: a network approach to understanding comorbidity. Multivariate Behavioral Research, 56:353–367, 2021.

[7] Jonas M. B. Haslbeck and Lourens J. Waldorp. How well do network models predict observations? on the importance of predictability in network models. Behavior Research Methods, 50:853–861, 2018.

[8] Michael N. Hallquist, Aidan G. C. Wright, and Peter C. M. Molenaar. Problems with centrality measures in psychopathology symptom networks: why network psychometrics cannot escape psychometric theory. Multivariate Behavioral Research, 56:199–223, 2021.

[9] Zachary P. Neal and Jennifer Watling Neal. Out of bounds? the boundary specification problem for centrality in psychological networks. Psychological Methods, 28:179–188, 2023.

[10] Zachary P. Neal et al. Critiques of network analysis of multivariate data in psychological science. Nature Reviews Methods Primers, 2:90, 2022.

[11] Gabriela Lunansky et al. Intervening on psychopathology networks: evaluating intervention targets through simulations. Methods, 204:29–37, 2022.

[12] Thomas R. Henry, Donald J. Robinaugh, and Eiko I. Fried. On the control of psychological networks. Psychometrika, 87:188–213, 2022.

[13] Tessa F. Blanken et al. Introducing network intervention analysis to investigate sequential, symptom-specific treatment effects: a demonstration in cooccurring insomnia and depression. Psychotherapy and Psychosomatics, 88:52–54, 2019.

[14] Judea Pearl. Causal inference in statistics: an overview. Statistics Surveys, 3:96–146, 2009.

[15] Julian Burger et al. Reporting standards for psychological network analyses in cross-sectional data. Psychological Methods, 28:806–824, 2023.

[16] Adela-Maria Isvoranu and Sacha Epskamp. Which estimation method to choose in network psychometrics? deriving guidelines for applied researchers. Psychological Methods, 28:925–946, 2023.

